# Non-invasive estimation of pressure drop across aortic coarctations: validation of 0D and 3D computational models with *in vivo* measurements

**DOI:** 10.1101/2023.09.05.23295066

**Authors:** Priya J. Nair, Martin R. Pfaller, Seraina A. Dual, Doff B. McElhinney, Daniel B. Ennis, Alison L. Marsden

## Abstract

**Purpose:** Blood pressure gradient (**Δ**P) across an aortic coarctation (CoA) is an important measurement to diagnose CoA severity and gauge treatment efficacy. Invasive cardiac catheterization is currently the gold-standard method for measuring blood pressure. The objective of this study was to evaluate the accuracy of **Δ**P estimates derived non-invasively using patient-specific 0D and 3D deformable wall simulations.

**Methods:** Medical imaging and routine clinical measurements were used to create patient-specific models of patients with CoA (N=17). 0D simulations were performed first and used to tune boundary conditions and initialize 3D simulations. **Δ**P across the CoA estimated using both 0D and 3D simulations were compared to invasive catheter-based pressure measurements for validation.

**Results:** The 0D simulations were extremely efficient (**∼**15 secs computation time) compared to 3D simulations (**∼**30 hrs computation time on a cluster). However, the 0D **Δ**P estimates, unsurprisingly, had larger mean errors when compared to catheterization than 3D estimates (12.1 **±** 9.9 mmHg vs 5.3 **±** 5.4 mmHg). In particular, the 0D model performance degraded in cases where the CoA was adjacent to a bifurcation. The 0D model classified patients with severe CoA requiring intervention (defined as **Δ**P **≥** 20 mmHg) with 76% accuracy and 3D simulations improved this to 88%.

**Conclusion:** Overall, a combined approach, using 0D models to efficiently tune and launch 3D models, offers the best combination of speed and accuracy for non-invasive classification of CoA severity.

## 1 Introduction

Coarctation of the aorta (CoA) is a congenital heart defect characterized by a constriction of the aorta, typically slightly distal to the origin of the left subclavian artery. It accounts for 6-8% of congenital heart defects [1], with an estimated incidence of 3 per 10,000 live births [2]. The narrowing of the aorta results in an increased resistance which causes proximal arterial hypertension and imposes a significant afterload on the left ventricle that can result in compensatory ventricular hypertrophy [3]. In the long term, it can result in complications such as stroke, early onset coronary artery disease, brain aneurysm, or aortic rupture due to the prolonged hypertension [4].

CoA results in a sudden drop in blood pressure (BP) across the aortic constriction. The BP gradient (ΔP) across the CoA and concomitantly elevated impedance can be alleviated using reconstructive surgical approaches or catheter-based stenting. For pre-operative evaluation of CoA severity, magnetic resonance imaging (MRI) or computed tomography (CT) are often used for anatomical assessment, which allows for geometric assessment of CoA, but these do not measure the functional consequences. Functional assessment is performed by measuring ΔP across the CoA. The current clinical indication of a severe CoA warranting corrective intervention is ΔP ≥ 20 mmHg at rest [5]. The gold-standard method for measuring ΔP is invasive cardiac catheterization.

Non-invasive clinical methods for estimating ΔP include Doppler echocardiography and, historically, estimating a difference in cuff BP measurements between the arms and legs. These non-invasive methods, however, are known to be less accurate than catheterization. Doppler echocardiography, with both the simplified and modified Bernoulli’s equation, overestimate ΔP by 41% on average [6]. The difference in BP between the upper and lower extremities has also been reported to be unreliable compared to the gold-standard, with an average error of 72% [7]. Therefore accurate clinical assessment of ΔP to determine CoA severity currently requires an invasive and costly catheter-based test. Improved non-invasive methods for functional assessment of CoA severity offer potential to avoid drawbacks and risks of invasive catheterization (including bleeding, infection, and exposing patients to radiation and contrast agents) and also lower the cost of patient care.

Computational fluid dynamics (CFD) is increasingly used for the non-invasive assessment of local hemodynamics in patients with cardiovascular disease. Anatomic and physiological data acquired non-invasively in the clinic can be used to generate patient-specific hemodynamic simulations. CFD simulations have been used extensively in pediatric cardiology for surgical planning and assessment of disease progression [8–12]. Previous studies have explored the use of CFD simulations to model hemodynamics in CoA, but these studies either had small sample sizes or assumed the aortic walls to be rigid [13–19]. While the wall deformability does not significantly alter the velocity field in arteries where the wall motion is small during the cardiac cycle [20], more significant differences do occur in large vessels like the aorta that undergo larger deformations. Rigid wall simulations also fail to capture important physiological phenomena such as wave propagation and reflections, which can have a significant impact on resulting solutions [21, 22]. However, 3D simulations, particularly those incorporating fluid-structure interaction (FSI), are computationally expensive (requiring ∼30 hours per simulation on a high-performance computing cluster), which currently limits their clinical use. The computational cost of 3D simulations is also amplified by the need for several repeated simulations for boundary condition tuning, and multiple cardiac cycles to reach periodic convergence.

Reduced-order models that are more computationally efficient (requiring only a few seconds per simulation on a local machine) therefore become an attractive tool that can be leveraged to accelerate 3D simulations. 0D models have no explicit spatial dependency and only depend on time, but are capable of predicting bulk quantities at different nodal locations in the cardiovascular model. 0D models are comprised of lumped-parameter elements such as resistors, capacitors, and inductors that connect to form an entire network. Previous work by Pfaller et al. demonstrated the robustness of 0D models by comparing to 3D results in 72 patient-specific cardiovascular models [23].

Herein, we explore the use of 0D and 3D simulations for efficient non-invasive estimation of ΔP in patients with CoA to guide clinical decision-making. The central goal of this study was to evaluate the accuracy of 3D simulations for ΔP estimation in patients with CoA and to demonstrate the utility of 0D simulations in accelerating 3D simulations.

## 2 Materials and Methods

In this section, we summarize the methods used to create patient-specific CoA models and perform 0D and 3D simulations. We also describe the analysis performed to compare these simulation results to *in vivo* pressures measured using a catheter.

### 2.1 Patient Data Acquisition

Under a protocol approved by the Stanford Institutional Review Board, patients with CoA from the Lucile Packard Children’s Hospital at Stanford were retrospectively identified. Patients with native and recurrent CoA were included. Inclusion criteria for the cohort required an imaging exam (4D-Flow MRI or CT) and invasive pressure measurements acquired via cardiac catheterization. Patients with aortic valve stenosis and/or extensive collaterals were excluded from this study. We obtained retrospective CT/4D-Flow MRI datasets for 17 patients with CoA (12 CT, 5 MRI, age = 29 ± 10.99 years, 13M / 4F). Invasive measurements of pressure via cardiac catheterization in the ascending aorta (AAo) and descending aorta (DAo) were obtained, in addition to cuff BP and heart rate. Informed consent was not required for this retrospective clinical data collection. The median time period between the imaging exam and the catheterization procedure was 96 days with a 95% confidence interval of [39, 203] days.

### 2.2 3D Model Construction

The CT or MRI magnitude images were imported into SimVascular (simvascular.org) [24]. Centerlines were drawn through the aorta and the branches arising from the aortic arch: brachiocephalic trunk, left common carotid artery, and left subclavian artery. 2D contours were drawn along the centerlines and then lofted to create the 3D geometry for each patient. The vessel wall was modeled as a thin membrane. The 3D blood volume and vessel wall were meshed using tetrahedral elements. We also incorporated a boundary layer mesh consisting of 3 layers to resolve the high velocity gradient at the wall. In the region of interest (at the CoA and the region immediately distal to it), we further refined the mesh to capture the high-velocity jet created by the stenosis as it travels through the vessel narrowing and subsequent post-stenotic dilation. The final meshes contained ∼2 million elements on average.

### 2.3 0D Model Construction

A 0D model typically consists of individual lumped-parameter elements (resistors, capacitors and inductors) that connect to form a lumped parameter network (LPN). The LPNs can be used to predict bulk hemodynamic quantities such as flow rate and spatially averaged pressure over time. 0D models are also referred to as circuit-analogy models because they are analogous to electrical circuits. Flow rate in the 0D model corresponds to current, just as pressure drop corresponds to voltage. Similarly, in the 0D model, resistance (*R*) captures the viscous nature of blood flow, capacitance (*C*) captures the elastic deformability of the vessel walls, and inductance (*L*) captures the inertia of blood flow. Drawing from the governing equations in electrical circuits, the dynamics of our 0D model elements are governed by:

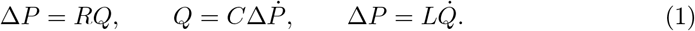

For a straight vessel with Poiseuille flow, the 0D elements can be described by:

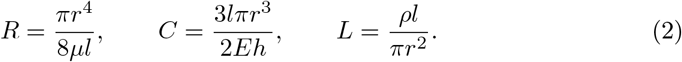

where *r* is the radius of the lumen, *μ* is the dynamic viscosity of the blood, *l* is the length of the vessel, *E* is the Young’s modulus of the vessel wall, *h* is the vessel wall thickness, and *ρ* is the density of blood. At the stenosis, due to the narrowing of the vessel and the resulting flow separation effects, the Poiseuille flow assumption underestimates the resistance. To account for this, an additional non-linear expansion-based resistance is added to the Poiseuille resistance (3) [25, 26]:

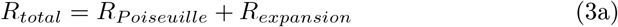

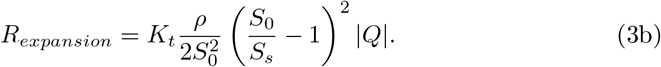

Here, *K*_*t*_ = 1.52 is a commonly used empirical correction factor [7, 25, 27]. *S*_0_ and *S*_*s*_ are the cross-sectional areas of the vessel lumen proximal to and at the location of the stenosis.

To create the 0D models, the vessel’s centerlines and cross-sectional areas were automatically extracted from the 3D models [23]. Vessel junctions (bifurcations) were then automatically identified and the centerline network was split into branches and junctions. Finally, stenoses were detected by sampling the cross-sectional area along the centerline of each branch and extracting the relative maxima (*S*_0_) and minima (*S*_*s*_) of the cross-sectional area. Each stenosed branch was then split into three segments: proximal, stenosis, and distal (with one 0D element per segment), to locate the stenosis at the correct location within a branch.

### 2.4 Boundary Conditions

We imposed a patient-specific inflow waveform (assuming a parabolic flow profile) as the inlet boundary condition (detailed in the subsequent section). A three-element Windkessel model (proximal resistance R_p_, capacitance C, distal resistance R_d_) was imposed at each of the outlets. The total peripheral resistance (R_total_) was determined using mean inflow and mean cuff BP (P_mean_), where P_mean_ was defined as:

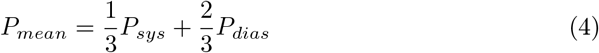

For patients with MRI and those with CT, total capacitance (C_total_) of 10^−5^ cm^5^/dyn was split to each of the branches proportional to the cross-sectional area of the branch.

We performed 0D simulations first and used them to fine-tune the prescribed boundary conditions. The value of C, along with R_p_/R_d_ ratio, were manually adjusted until the calculated pressures matched the patient’s systolic (*P*_*sys*_) and diastolic (*P*_*dias*_) cuff BP measurements within 5 mmHg. Boundary condition tuning typically required about 5-7 iterations of the 0D simulation (less than 2 minutes of total computation time) and removed the need for time-consuming repeated 3D simulations. The specific tuning process differed for patients who had an MRI versus those who had a CT. The details of the tuning process for each of these subsets are described below and outlined in Figure 1 and Table 1.

**Fig. 1.**
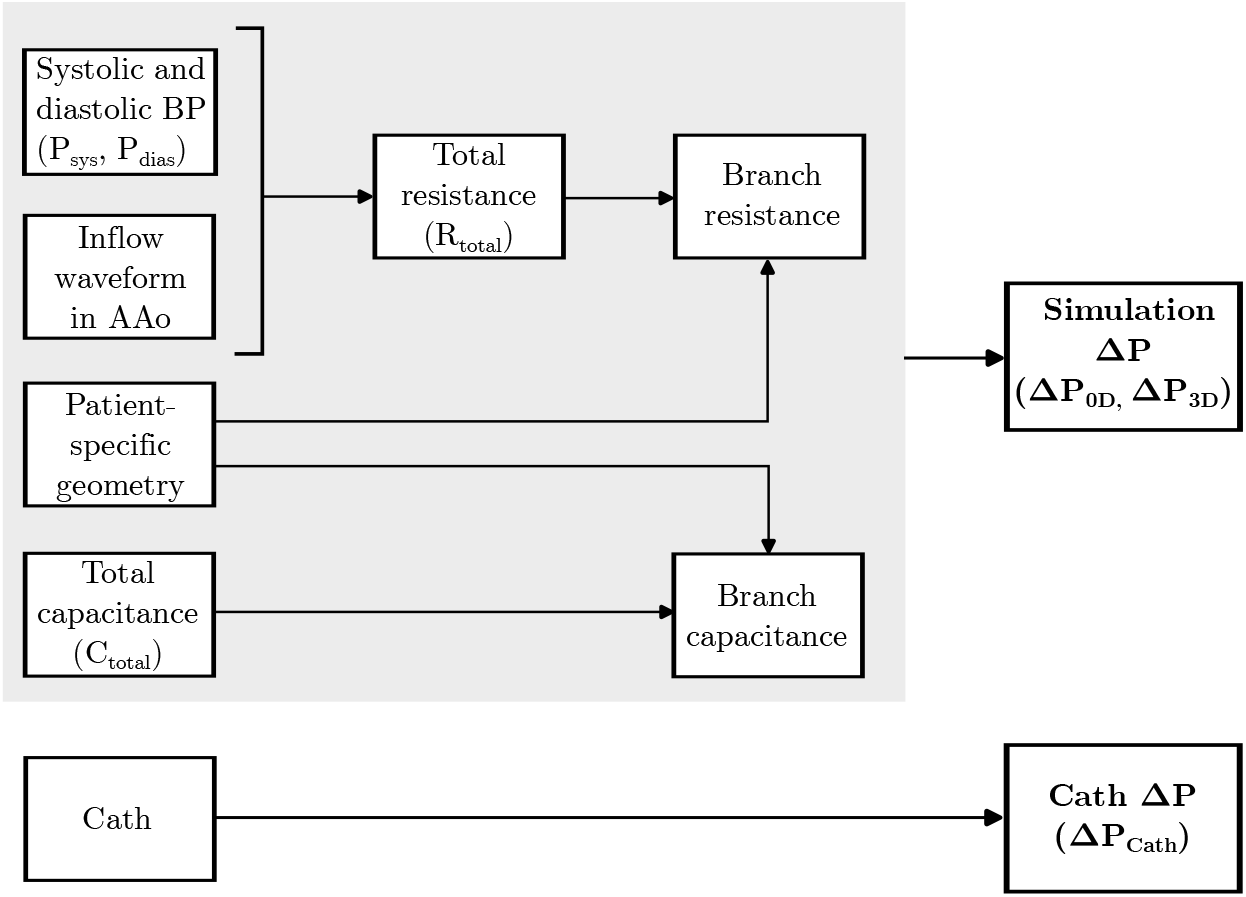
Strategy for personalization of patient-specific models.

**Table 1.** Sources for each parameter used to personalize patient-specific 0D and 3D models for patients with MRI and patients with CT.

| Parameter | Source |  |
| --- | --- | --- |
|  | MRI (n=5) | CT (n=12) |
| Patient-specific geometry | Magnitude images | CT images |
| Inflow waveform in AAo | 4D-Flow MRI | Averaged waveform scaled using CO and HR |
| Systolic and diastolic BP | Cuff measurement | Cuff measurement |
| Branch resistance | $R_{total}$ split by flow split determined from 4D-Flow MRI | $R_{total}$ split by cross-sectional area |
| Branch capacitance | $C_{total}$ split by cross-sectional area | $C_{total}$ split by cross-sectional area |

#### 2.4.1 Patients with 4D-Flow MRI

We derived inflow waveforms (averaged over the vessel cross-sectional area) from the patients’ 4D-Flow MRI exams. Eddy current correction was applied using a machine learning-based correction tool available in Arterys (Arterys, San Francisco, USA), followed by further manual correction. Contours of the inlet (just past the aortic sinus) and outlets (at the vessel entries) were manually drawn on the eddy current-corrected images. From the 4D-Flow MRI dataset we quantified 2D time-resolved flow averaged over the cross-sectional area. The patient-specific temporally varying flow profile obtained from 4D-Flow MRI was prescribed to the inlet of the 0D and 3D models. The resistance R_p_ + R_d_ for each of the branches was calculated by distributing R_total_ to each of the branches as inversely proportional to the flow splits determined from 4D-Flow MRI.

#### 2.4.2 Patients with CT

We generated a generalized inflow waveform by averaging the MRI-derived inflow waveforms of seven CoA patients in the Vascular Model Repository (vascularmodel.org). These patients were unrelated to the patient cohort used for this study. The systolic portions of the waveforms were averaged and scaled to be 1/3^rd^ of the average cardiac cycle. Similarly, the diastolic portions were scaled to be 2/3^rd^ of the cardiac cycle. We then scaled the generalized waveform to match the patient’s cardiac output (measured during catheterization using the Fick’s method) and heart rate. This patient-specific temporally varying flow profile was prescribed to the inlet of the 0D and 3D models (assuming parabolic flow). The resistance R_p_ + R_d_ for each of the branches was then calculated by distributing R_total_ to each of the branches as inversely proportional to the cross-sectional area of that branch.

### 2.5 Simulations

After efficiently optimizing the boundary conditions, for each patient model we first ran the 0D simulation for 10 cardiac cycles to ensure the pressures reached periodic convergence. We then projected the results from the final cardiac cycle of the 0D simulation onto the 3D mesh. This projection was used to initialize the 3D simulation [28], allowing the 3D simulation to converge to a steady solution with lower computation time than it would without initialization. (Figure 2). The simulation results without initialization do not reach a converged state even after 10 cardiac cycles, but the steady state initialized simulation and the 0D initialized simulation achieved periodic convergence in 5 and 6 cardiac cycles, respectively in one representative case. Convergence was defined as an asymptotic error in pressure ≤ 1% [28]. The computational cost of each of these initializations varied greatly, with the 3D steady flow rigid simulation requiring 2.5 hours on a high-performance computing cluster, whereas the 0D simulation required negligible computation time (15 seconds) on a local computer for the same number of cardiac cycles.

**Fig. 2.**
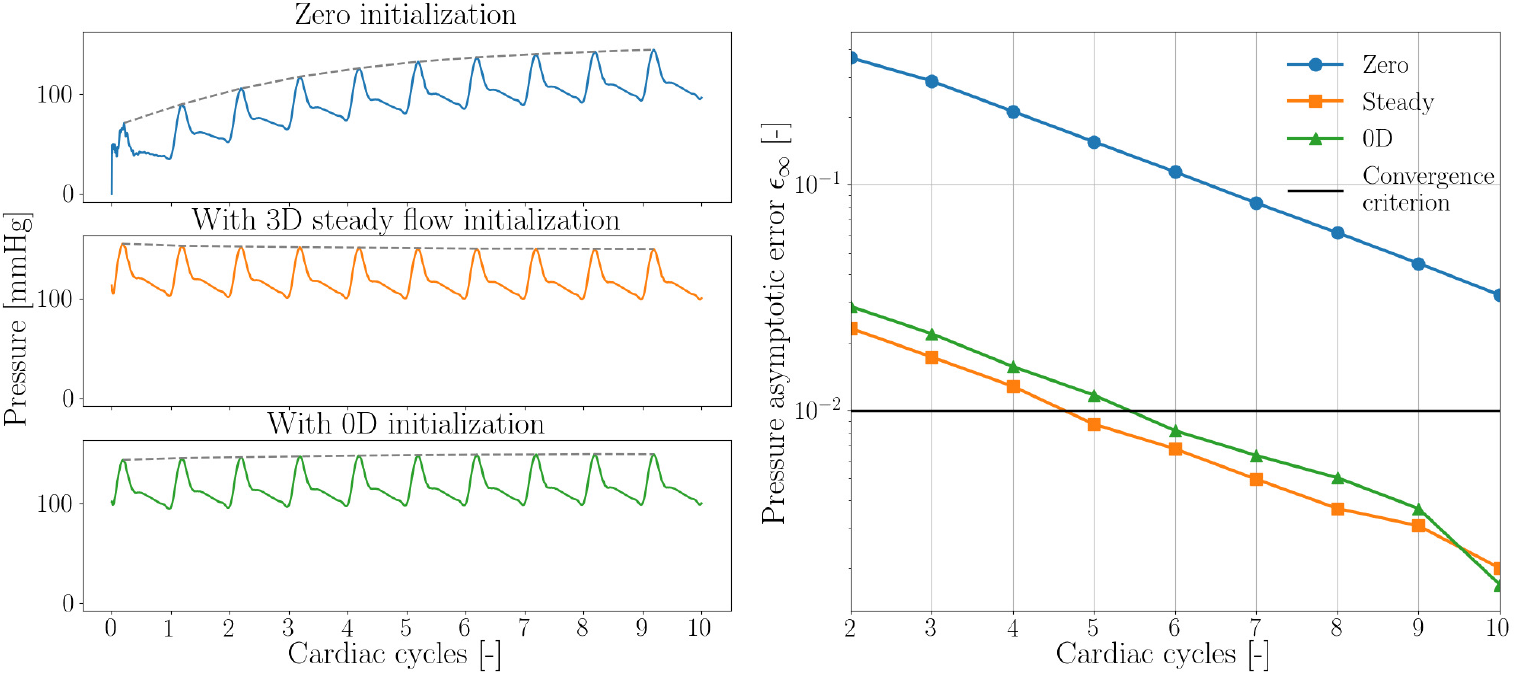
Convergence of aortic pressure to a steady solution without initialization (blue), initialization using results from a 3D steady flow rigid wall simulation (orange), and using results from a 0D simulation (green).

We performed 3D simulations using the coupled momentum method (CMM) with svSolver, SimVascular’s finite element solver for fluid-structure interaction between an incompressible, Newtonian fluid and a linear elastic membrane for the vascular wall [24, 29]. The solver has been validated in prior studies [30, 31]. The same boundary conditions used in the 0D simulation were prescribed for the 3D simulation. The Young’s modulus of the aortic wall was prescribed to be 3x10^6^ dyn/cm^2^, based on previously reported values of stiffness in a human aorta with CoA [15]. The thickness of the wall was assumed to be 10% of the diameter of the vessel. A Poisson’s ratio 0.5, shear constant 0.8333, and density 1 g/cm^3^ were used to further define the vessel wall material properties. The fluid was prescribed to have density 1.06 g/cm^3^ and viscosity 0.04 Poise to match the properties of blood. 3D simulations were run for 10 cardiac cycles to ensure that the pressures reach periodic convergence; only the final cardiac cycle was analyzed.

We measured the pressure averaged over the cross-sectional area in the AAo and DAo at peak systole in the 0D and 3D models. Pressure in the AAo was defined as the mean of the pressure measured in the last 20% of the length of the AAo since the exact catheter location was unknown. Pressure in the DAo was defined as the centerline pressure measured at a point in the DAo two vertebral spaces above the diaphragm to match the typical catheter location. ΔP was calculated as the difference between the pressures in the AAo and DAo. ΔP_0D_ and ΔP_3D_ were compared to ΔP_Cath_.

### 2.6 Statistical Analysis

We used paired Student’s t-test to compare ΔP_0D_ and ΔP_3D_ to ΔP_Cath_. A p-value of was considered significant. The agreement of ΔP_0D_ and ΔP_3D_ with ΔP_Cath_ was characterized with Bland-Altman plots. We performed bootstrapping to estimate the population mean and standard deviation of the population based on the sample of 17 patients with CoA.

## 3 Results

A total of 17 patients with CoA were included in the study (Table 2). The variety of aorta geometries, locations of stenosis and range of blood pressures represented in this patient cohort are depicted in Figure 3. The patient cohort can be divided into two categories - ones where the CoA occurs in a junction region and ones where the CoA occurs in a vessel (“non-junction”) region.

**Table 2.**
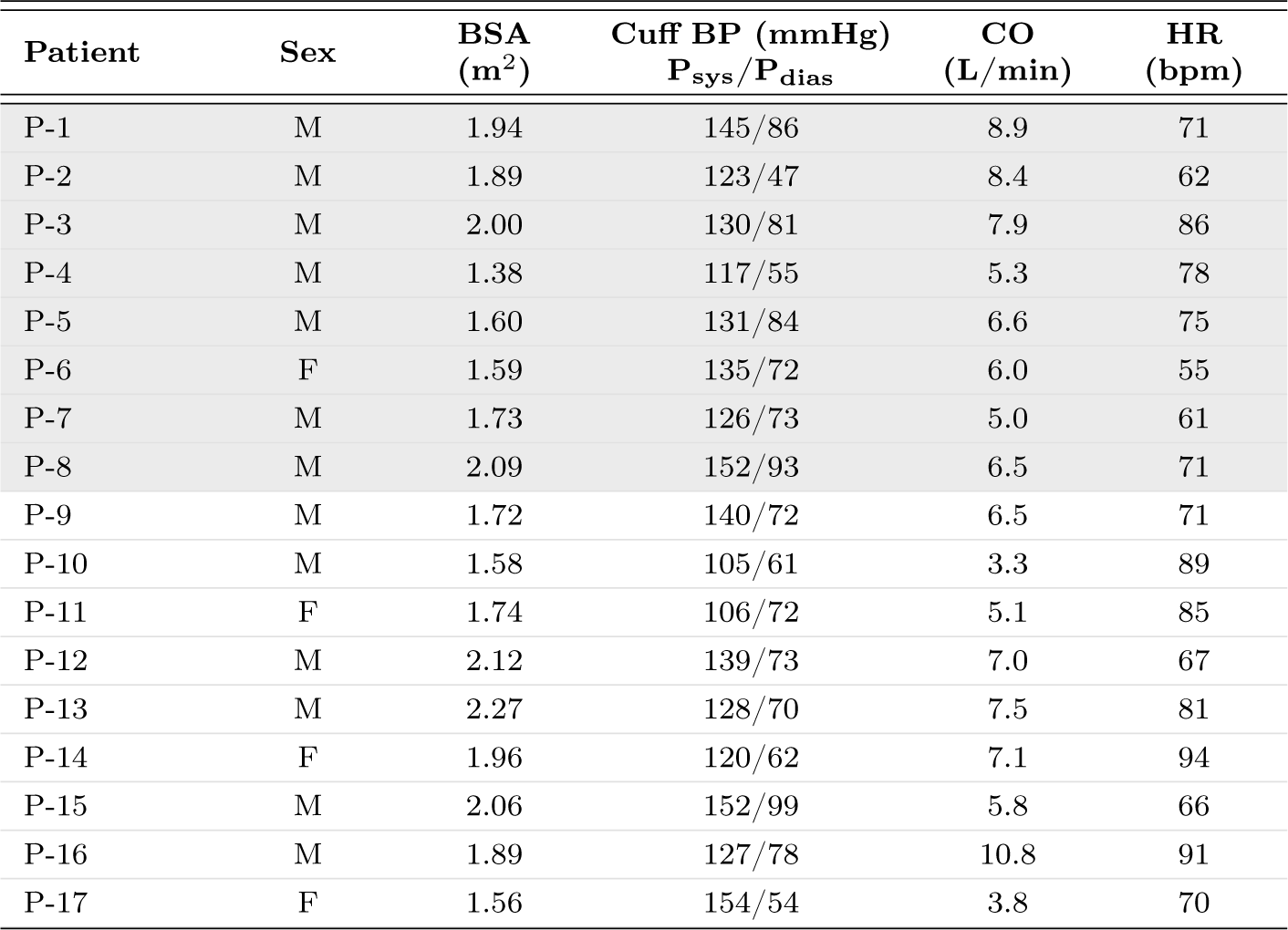
Patient characteristics. Patients with junction CoA are shaded in gray. BSA: Body surface area, CO: Cardiac output, HR: Heart rate.

| Patient | Sex | BSA<br>(m<sup>2</sup>) | Cuff BP (mmHg)<br>P<sub>sys</sub>/P<sub>dias</sub> | CO<br>(L/min) | HR<br>(bpm) |
| --- | --- | --- | --- | --- | --- |
| P-1 | M | 1.94 | 145/86 | 8.9 | 71 |
| P-2 | M | 1.89 | 123/47 | 8.4 | 62 |
| P-3 | M | 2.00 | 130/81 | 7.9 | 86 |
| P-4 | M | 1.38 | 117/55 | 5.3 | 78 |
| P-5 | M | 1.60 | 131/84 | 6.6 | 75 |
| P-6 | F | 1.59 | 135/72 | 6.0 | 55 |
| P-7 | M | 1.73 | 126/73 | 5.0 | 61 |
| P-8 | M | 2.09 | 152/93 | 6.5 | 71 |
| P-9 | M | 1.72 | 140/72 | 6.5 | 71 |
| P-10 | M | 1.58 | 105/61 | 3.3 | 89 |
| P-11 | F | 1.74 | 106/72 | 5.1 | 85 |
| P-12 | M | 2.12 | 139/73 | 7.0 | 67 |
| P-13 | M | 2.27 | 128/70 | 7.5 | 81 |
| P-14 | F | 1.96 | 120/62 | 7.1 | 94 |
| P-15 | M | 2.06 | 152/99 | 5.8 | 66 |
| P-16 | M | 1.89 | 127/78 | 10.8 | 91 |
| P-17 | F | 1.56 | 154/54 | 3.8 | 70 |

**Fig. 3.**
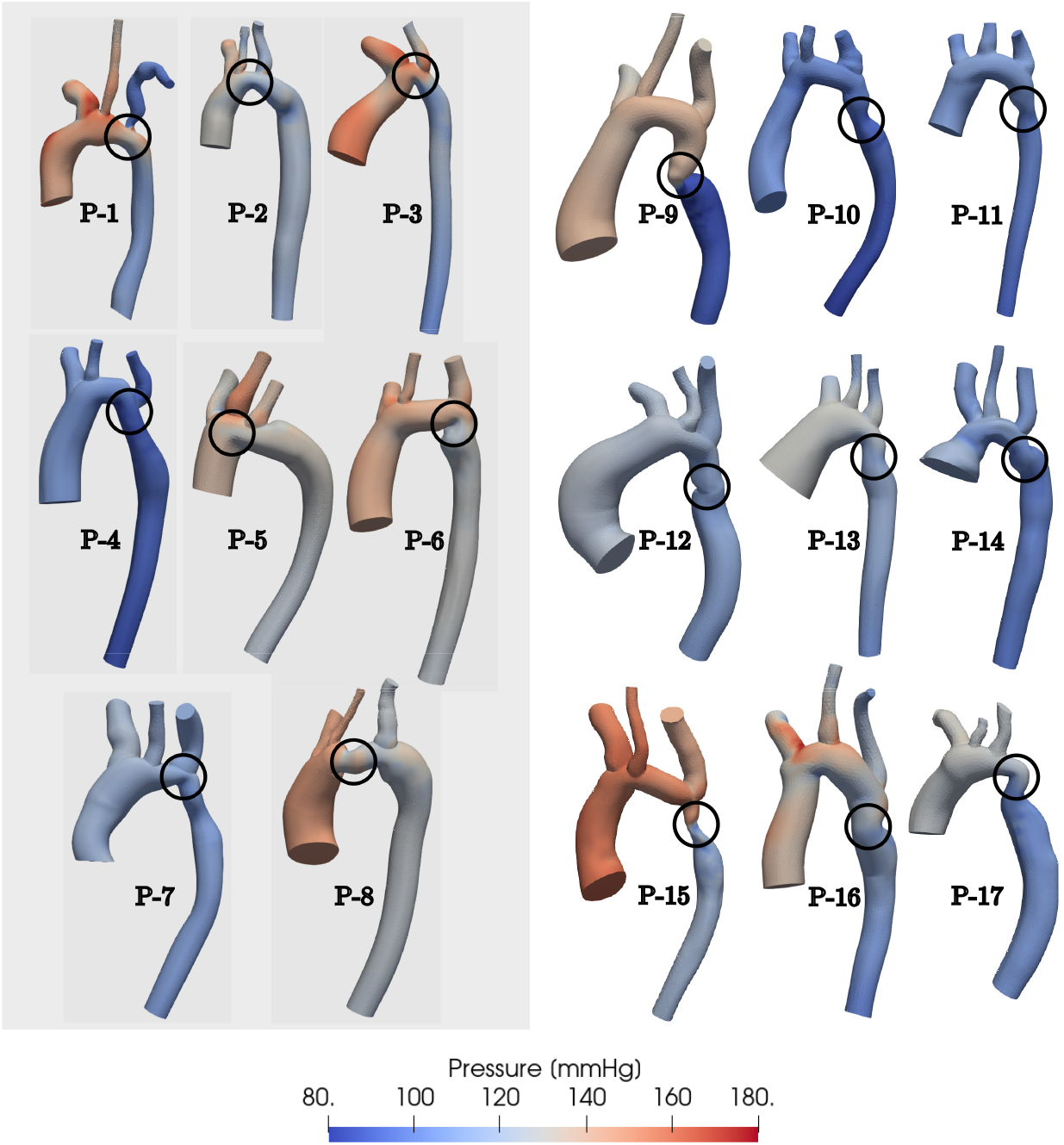
3D pressure distributions in N = 17 patients with CoA at peak systole. Patients with junction CoA are shaded in gray (left).

The 0D simulations were far more efficient than the 3D simulations. Each 0D simulation had a computation time of approximately 15 seconds on a local computer. While the use of 0D simulations to tune boundary conditions and initialize the 3D simulations helped to reduce some of the computational cost of running 3D simulations, each 3D simulation still required almost 30 hours of computation time on a high-performance computing cluster. To compare the accuracy of 0D and 3D simulation results against clinical data, the specific measurements of ΔP using 0D simulations, 3D simulations, and catheterization for the entire patient cohort are reported in Table 3. The difference between ΔP_3D_ and ΔP_Cath_ had a lower root mean square (RMS) value of 5.3 mmHg and a smaller standard deviation (SD) of 5.4 mmHg compared to the difference between ΔP_0D_ and ΔP_Cath_ which had an RMS of 12.1 mmHg and SD of 9.9 mmHg. Figure 4 depicts the results of 0D and 3D simulations in two representative patients, one each from the junction and non-junction CoA subsets. The accuracy of the 0D models’ pressure estimates vary for each of these categories. As depicted in Figure 4, the 0D model fails to provide an accurate estimate of ΔP across the CoA when the stenosis is in a junction region (left) due to the current 0D model generation process, a source of error that is discussed later. 0D models can, however, produce reasonably accurate ΔP estimates compared to 3D simulations and catheter-measured pressures when the stenosis is in a non-junction region. This difference in accuracy can be visualized in more detail in Figure 5 which depicts the pressure along the centerline in the same two representative patients. In both patients, the 3D simulations (solid line) accurately captures ΔP across the CoA. While the 0D model (dotted line) closely matches 3D simulation results in P-9 (“non-junction” CoA), it fails to capture ΔP across the CoA in P-1 (“junction” CoA).

**Table 3.** Comparison of pressure drops (ΔP AAo-DAo) from 0D and 3D simulations with catheter-derived measurements.

| Patient | $\Delta P_{0D}$ | $\Delta P_{3D}$ | $\Delta P_{Cath}$ |
| --- | --- | --- | --- |
|  | [mmHg] |  |  |
| P-1 | 8 | 31 | 30 |
| P-2 | 6 | 12 | 10 |
| P-3 | 16 | 22 | 18 |
| P-4 | 9 | 13 | 10 |
| P-5 | 2 | 11 | 15 |
| P-6 | 3 | 13 | 18 |
| P-7 | 4 | 6 | 24 |
| P-8 | 1 | 22 | 20 |
| P-9 | 49 | 46 | 43 |
| P-10 | 14 | 12 | 10 |
| P-11 | 11 | 7 | 10 |
| P-12 | 2 | 9 | 7 |
| P-13 | 10 | 14 | 19 |
| P-14 | 8 | 10 | 10 |
| P-15 | 40 | 37 | 36 |
| P-16 | 3 | 26 | 28 |
| P-17 | 9 | 15 | 11 |

**Fig. 4.**
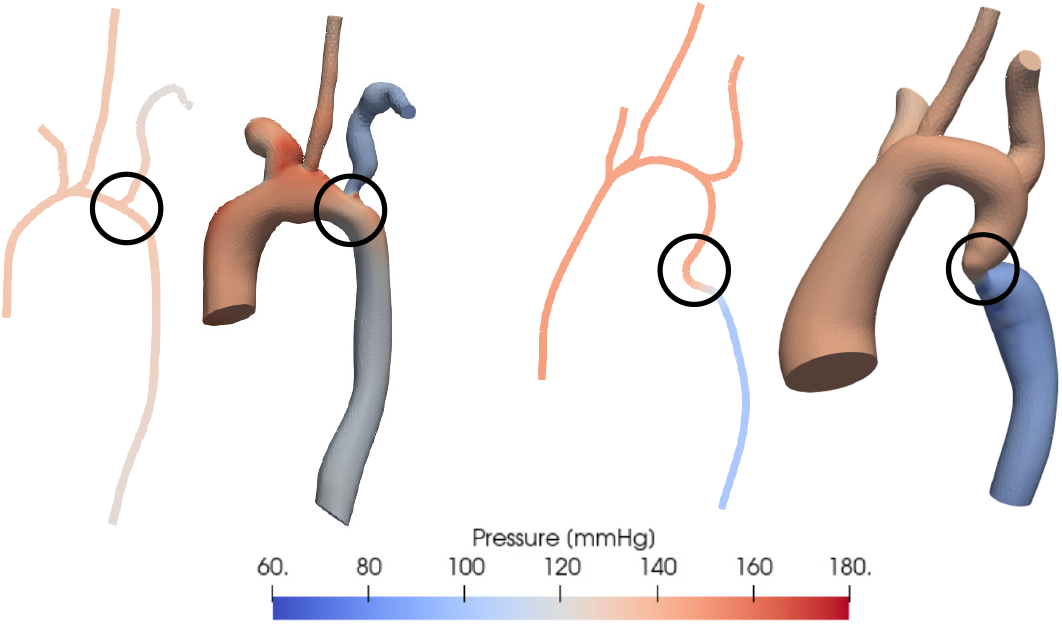
Comparison of pressure distribution estimated using 0D (left) and 3D (right) simulations in 2 representative patients with vessel narrowing at different locations - at a vessel junction and in the descending aorta. The location of the coarctation is circled in black.

**Fig. 5.**
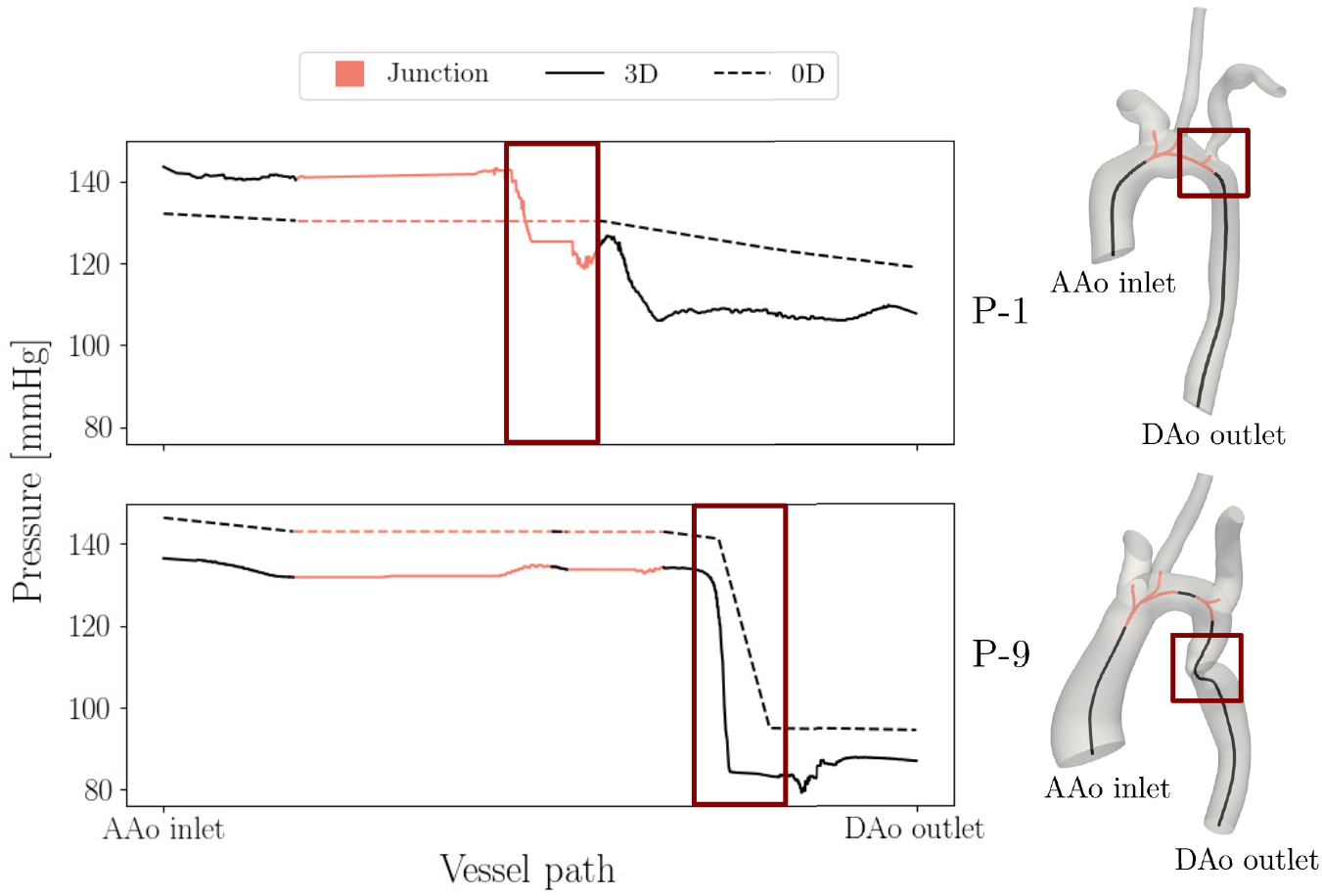
Pressure over the normalized vessel path along the aorta estimated using 0D (dotted) and 3D (solid) simulations at peak systole. Results are depicted for two representative patients with CoA at different locations (red box) - at a vessel junction (top) and in the DAo (bottom). Junctions regions are depicted in pink.

The measurements of ΔP_0D_ depicted in Figure 6 show a larger deviation from ΔP_Cath_ for patients with CoA in the junction, while ΔP_3D_ matches ΔP_Cath_ very closely in most cases.

**Fig. 6.**
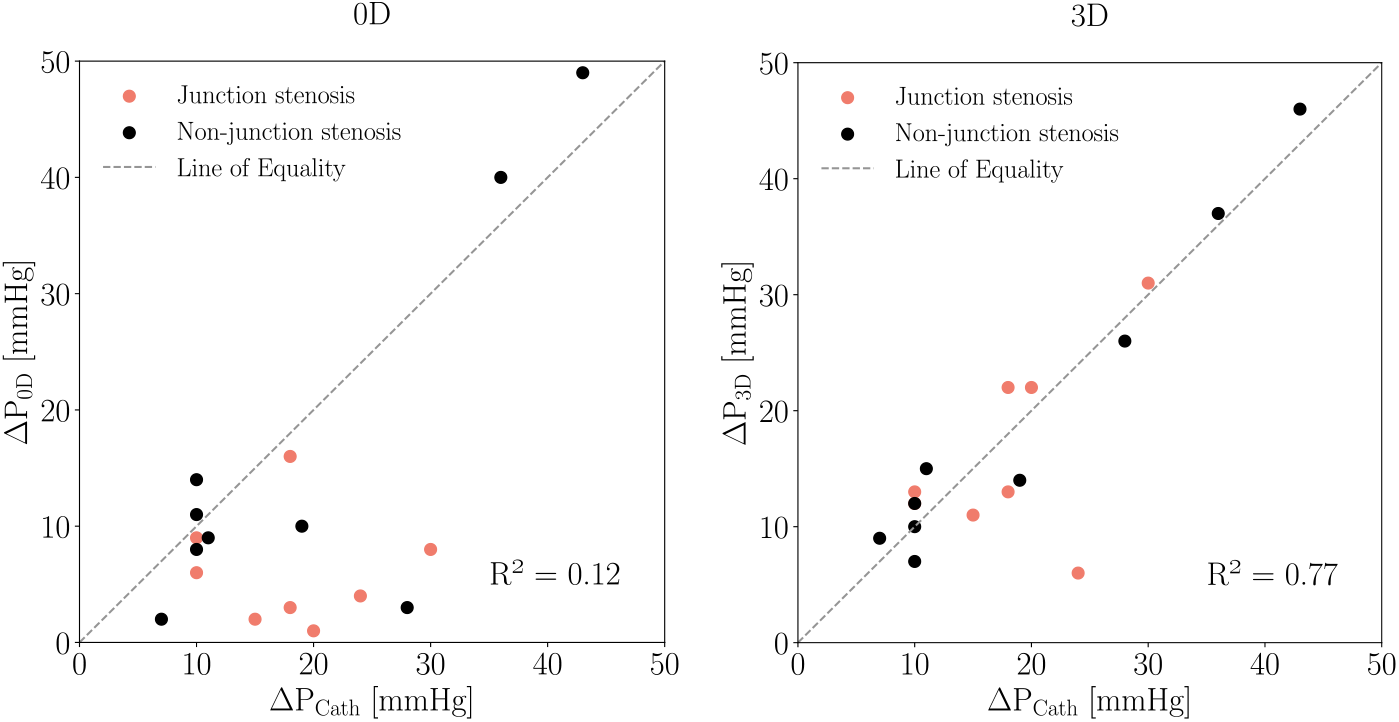
Comparison of CoA pressure drop estimated using 0D (left) and 3D (right) simulations with catheter-derived pressure measurements. Dotted line indicates no error between simulation results and catheter measurements.

The agreement between simulation and catheter-derived ΔP estimates was evaluated using Bland-Altman analysis shown in Figure 7. Both ΔP_0D_ and ΔP_3D_ resulted in average underestimation of ΔP_Cath_ that was larger in the case of ΔP_0D_. The bias (mean of differences) was higher for ΔP_0D_ (−7.29 mmHg) than for ΔP_3D_ (−0.76 mmHg). The corresponding limits of agreement were ± 18.08 mmHg and ± 9.39 mmHg for ΔP_0D_ and ΔP_3D_, respectively. For the cases that specifically had a “non-junction” CoA, both the 0D and the 3D simulations provided closer estimates to ΔP_Cath_ than for “junction” CoA cases. The bias of ΔP_0D_ was -3.1 ± 15.6 mmHg versus -12 ± 14.4 mmHg and for ΔP_3D_ was -0.23 ± 5.23 mmHg versus -1.88 ± 11.6 mmHg for “junction” versus “non-junction” CoA.

**Fig. 7.**
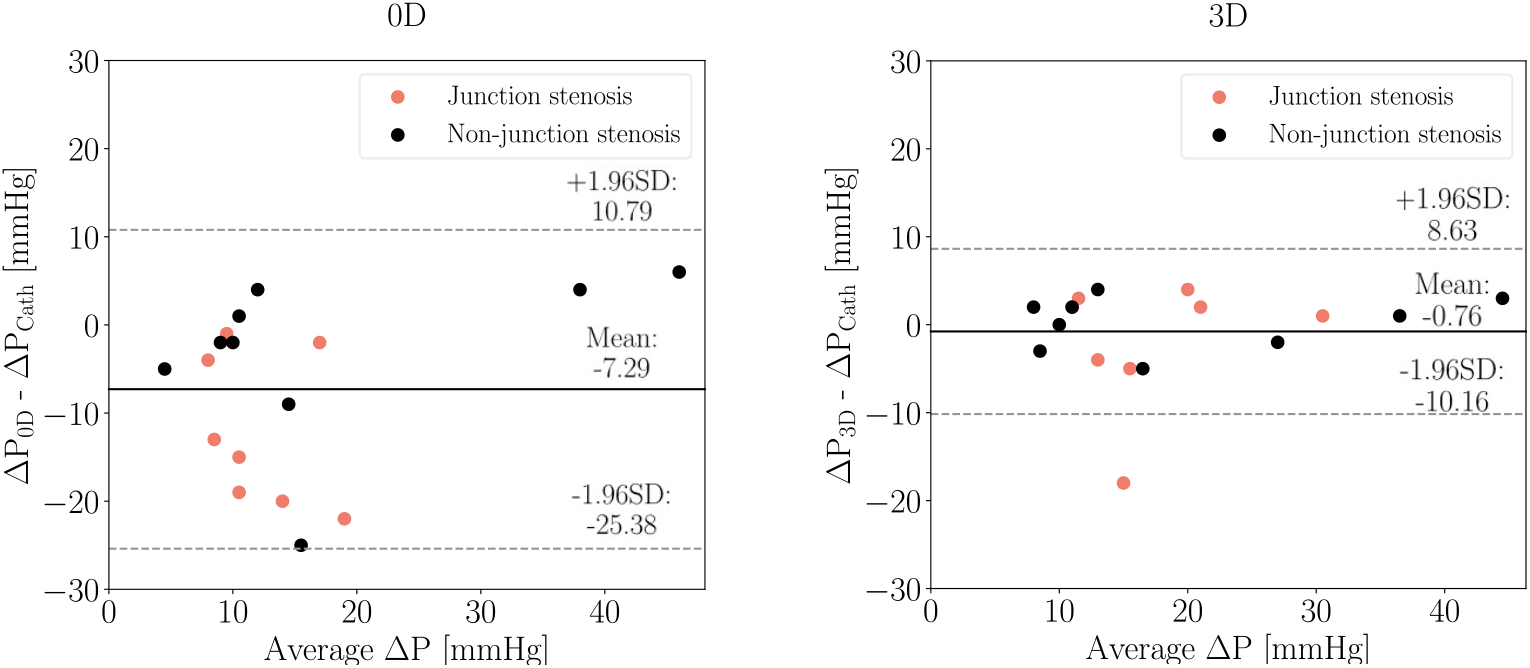
Bland-Altman plots of catheter-derived pressure gradient and estimates from 0D (left) and 3D (right) simulations. The solid lines represent the mean difference between simulation and catheter pressure gradient estimates, and the paired dotted lines correspond to the 95% limits of agreement.

In the clinical setting, a ΔP cutoff of 20 mmHg at rest across the CoA is used to guide the decision to intervene. ΔP < 20 mmHg is classified as a “mild” CoA and ΔP ≥ 20 mmHg is considered a “severe” CoA likely needing corrective intervention. Using this cutoff, ΔP_0D_ and ΔP_3D_ would result in the right treatment recommendation in 76% and 88% of cases, respectively (Figure 8). If a severe CoA is considered “positive”, the 0D estimate has a sensitivity of 33.3% and specificity of 100%. The 3D estimate has a sensitivity of 83.3% and specificity of 90.9%. In junction CoA cases, the 0D estimate has sensitivity and specificity of 0% and 100% respectively while the 3D estimate has sensitivity and specificity of 66% and 80% respectively. In non-junction CoA cases, the 0D estimate has sensitivity and specificity of 66% and 100% respectively while the 3D estimate has sensitivity and specificity of 100% and 100% respectively.

**Fig. 8.**
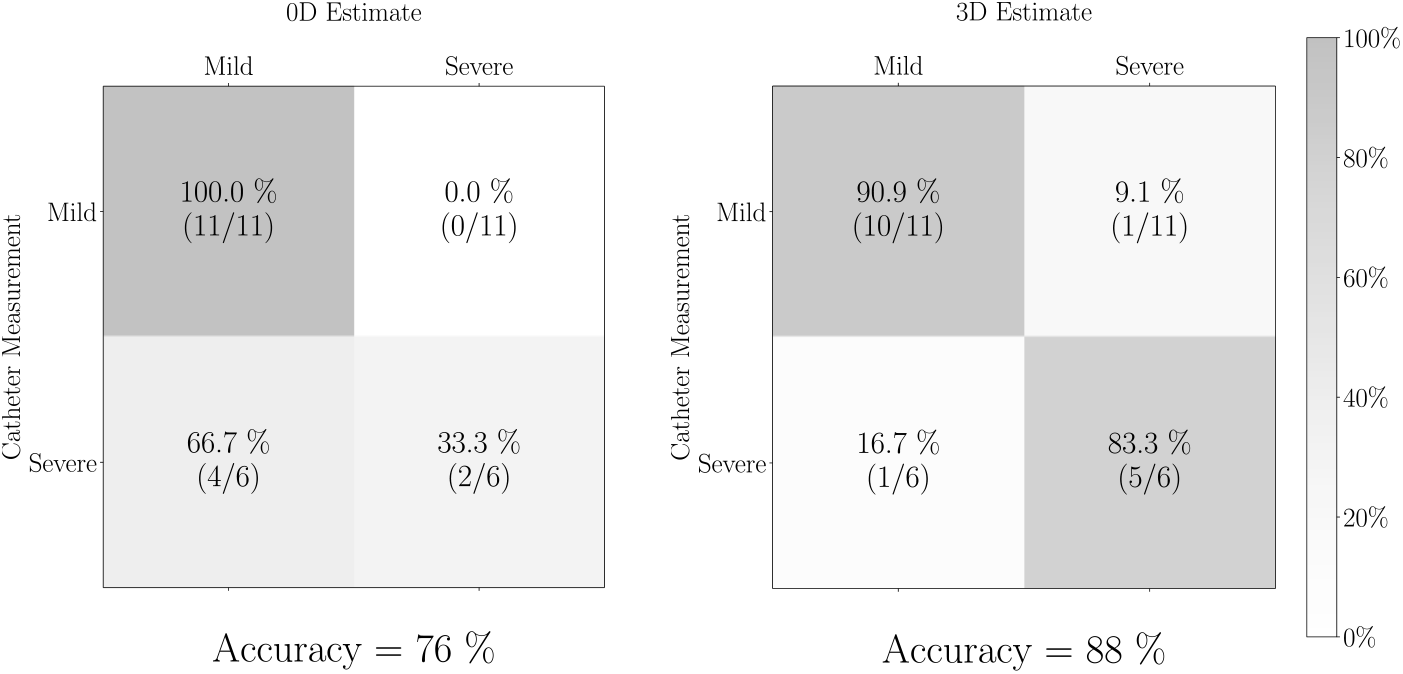
Confusion matrix for severity of CoA as determined by ΔP_Cath_ and ΔP_0D_ (left), ΔP_Cath_ and ΔP_3D_ (right). “Mild”: ΔP < 20 mmHg, “Severe”: ΔP ≥ 20 mmHg.

Paired Student’s t-test between ΔP_0D_ and ΔP_Cath_ had a p-value < 0.001 indicating that ΔP_0D_ was not always a good estimate of clinical catheter-derived pressures. The t-test between ΔP_3D_ and ΔP_Cath_ had p-value = 0.565, indicating that the null hypothesis was not rejected and that ΔP_3D_ provides non-invasive estimates of catheter pressures that are not significantly different.

## 4 Discussion

Invasive catheterization is the current gold standard for treatment decisions in patients with CoA. However, it is a costly procedure which is not risk-free, and thus there is a need for accurate non-invasive alternatives for estimation of ΔP. This study addresses this need and validates a CFD-based method for non-invasive estimation of aortic BP gradient in patient-specific models of CoA. 0D models (constructed automatically from 3D models) were used to efficiently tune boundary conditions, to initialize 3D simulations, and to compute flow and pressure in patient-specific models of CoA for comparison with full fidelity simulations.

ΔP predicted from 0D and 3D simulation validated using invasive catheter measurements had smaller percentage errors with catheterization than Doppler and arm-leg cuff BP measurements. 3D models estimated ΔP within 5 mmHg in most cases and had an average error of 12%. The 0D model had an average error of 30%, but was also a good estimator of catheter-derived pressure gradients in cases where the CoA was in a non-junction region.

The significantly lower computational time of the 0D models make them more feasible to use in a clinical setting in scenarios where the CoA is not located at a vessel junction. While 0D simulations are computationally efficient, the simplified nature of these models and lack of features such as wave propagation affects their accuracy. Unsurprisingly, the results of our study confirmed that the 0D model is not a sufficiently accurate standalone model for estimating ΔP in patients with CoA. One potential source of error in 0D models is the automatic stenosis detection. The vessel cross-sectional areas *S*_0_ and *S*_*s*_ (3) are crucial to accurately determine the resistance in the stenosis. In our framework, *S*_0_ and *S*_*s*_ are determined automatically and can give rise to errors when comparing the results of 0D and 3D simulations. Additionally, due to the complexity of solving for pressures and flows at junctions, the current 0D solver implements mass conservation at junction regions and assumes the pressure is constant between the inlets and outlets of the junction. Pressure drops in a junction, which often result from nonlinear effects such as flow separation, are therefore not accurately resolved. With future improvements to 0D models, they have the potential to become more accurate. Therefore, while 0D simulations cannot currently replace 3D simulations, they can be used to speed up the 3D model personalization process. They can be used as an efficient surrogate to iteratively tune boundary conditions within a couple of minutes. 3D simulations initialized with 0D results also converge in fewer cycles than they would without any initialization, thereby decreasing the computational cost and time of running a 3D CFD simulation.

This study has several limitations. Patients with CoA combined with aortic valve stenosis were excluded from the study given the lack of information to recreate the spatial inflow pattern from CT images in simulations which would significantly affect ΔP estimates downstream. Patients who had extensive collaterals were also excluded because of the lack of image resolution to segment these small vessels and quantify flow through them. Second, the same Young’s modulus obtained from literature was used for the vessel walls for all patients in this cohort, making the CFD results prone to error in the case of a mismatch; this information is not currently available on a patient specific basis. Third, while inflow measurements from the day of the imaging exam were used to inform the models for patients with MRI, pressure estimates derived from these models were compared to pressures measured on the day of catheterization (which was on average 80 days after the MRI exam). Any differences in the patient’s hemodynamic state between the MRI exam and the catheterization procedure would therefore contribute to errors.

For patients who only had a CT, their inflow waveform was an averaged waveform scaled to match the patient’s CO and HR. The CO used for this was estimated using the Fick’s method during the catheterization procedure. Therefore, for patients with CT analysis, this study’s proposed method is not entirely non-invasive. In the future, CO measurements from echocardiography or MRI could be used to make the method completely non-invasive. The patient-specific models and simulations framework described in this study can also be extended to study the impact of exercise on hemodynamics, particularly ΔP, in CoA patients. This metric cannot reliably be measured in the clinic. Future studies could also aim to address the inability of the 0D models to detect pressure drops at junctions. Machine learning models trained on hundreds of cardiovascular models in the Vascular Model Repository (vascularmodel.org) to better estimate pressure loss across junctions are currently in development and will be incorporated into subsequent iterations of the 0D model.

In conclusion, we have shown the capability of a combined 0D-3D simulation approach that accurately recapitulates invasive cardiac catheterization measurements and can guide patient-specific treatment planning in patients with CoA. Future work should assess the predictive accuracy of our methods in a prospective clinical study with a larger cohort.

## Data Availability

All data produced in the study will be publicly available in the Vascular Model Repository (vascularmodel.org)

http://vascularmodel.org/

## Acknowledgements

The authors thank the Stanford Research Computing Center for computational resources (Sherlock HPC cluster). This work is supported by the Vera Moulton Wall Center for Pulmonary Vascular Disease at Stanford University. PJN is supported by the National Science Foundation Graduate Research Fellowship (DGE-1656518) and MRP is supported by the Stanford Maternal and Child Health Research Institute and the National Institutes of Health (K99HL161313).

## Statements and Declarations

### Competing interests

The authors have no relevant financial or non-financial interests to disclose.

### Availability of data and materials

The datasets generated during this study will be available in the Vascular Model Repository (vascularmodel.org).

### Authors’ contributions

P.J.N designed the study, performed the computational simulations and analysis, and drafted the manuscript; M.R.P implemented the 0D model formulation in SimVascular and supported the computational simulations and analysis; S.A.D. extracted hemodynamic parameters from patient scans and supported the computational analysis; D.B.M provided patient data and clinical insights used to conceptualize this study; D.B.E provided overall supervision and advice to the research; A.L.M conceptualized the study and provided overall supervision to the research work. All authors have reviewed the final manuscript.

## Notes

### Competing Interest Statement

The authors have declared no competing interest.

### Author Declarations

All patient data used in this study was acquired as part of the usual clinical care for the patients. The IRB of Stanford University gave ethical approval for this work.

